# Early Repolarization Pattern is Associated with Schizophrenia: A Single Center Experience in Japan

**DOI:** 10.1101/2020.07.16.20155838

**Authors:** Hiroshi Kameyama, Kenichi Sugimoto, Keisuke Inamura, Kyoko Itoh, Fumitoshi Kodaka, Yuki Matsuda, Kazutaka Nukariya, Tomohiro Kato, Masahiro Shigeta

**Affiliations:** Department of Psychiatry, The Jikei University School of Medicine; Department of Cardiology, The Jikei University School of Medicine; Center for Predicitive Medicine, The Jikei University Hospital

**Author notes:** **Correspondence to: Name:** Hiroshi Kameyama, **Address:** Department of Psychiatry, The Jikei University School of Medicine, 3-19-18, Nishi- Shimbashi, Minato-Ku, Tokyo 105-8471, Japan. **Conflict of Interest:** none.

**Keywords:** Cardiovascular disease, Early repolarization pattern, Schizophrenia, Electrocardiogram

## Abstract

**Introduction:** Recent studies have shown a high frequency of abnormal electrocardiograms in patients with schizophrenia. The objective of this study was to associate schizophrenia diagnoses with early repolarization patterns in a sample of hospitalized patients from a single hospital in Japan.

**Methods:** We conducted a retrospective age, sex and coronary risk factors matched case-control study on 85 patients with schizophrenia and 89 controls from medical checkups. First, we compared the presence of early repolarization patterns in both groups. Secondly, we elucidated an association between the presence of an early repolarization pattern and clinical findings in the patients’ groups. We also evaluated J-point elevation patterns.

**Results:** As a result, we found that both early repolarization patterns and J-point elevation patterns observed were significantly higher in the schizophrenic group than in the matched control group (early repolarization pattern 23;6 P < 0.001; J-point elevation pattern 34:12; P = 0.001). After multivariable logistic regression among the patients and controls, schizophrenia was the independent predictor for early repolarization pattern (P = 0.001) and J-point elevation (P < 0.001). Among the patients, the independent predictor for early repolarization pattern was psychiatric family history (P = 0.006), while older age (P = 0.038) and psychiatric family history (P = 0.014) were predictors for J-point elevation patterns.

**Conclusion:** These findings suggest that an association between early repolarization pattern or J-point elevation pattern and schizophrenia in a single Japanese center.

## Introduction

Schizophrenia, which is characterized by hallucinations and delusions, is a common familial psychiatric disorder and is prevalent in about 1% of the general population [1]. Despite intensive study, its molecular etiology remains enigmatic [2]. The outcome of schizophrenia has improved significantly with the administration of antipsychotics based on the dopamine hypothesis [3].

However, about 30% of patients with schizophrenia did not responses to conventional therapy, experiencing relapses and a deterioration in their symptoms, and requiring hospitalization [4]. In these refractory cases, there is a different pathological mechanism which does not match the dopamine hypothesis [5]. A biological indicator for these patients is also required. Many biological indicators have been studied for central nervous disorders including mental disorders, with an association between abnormalities in electrocardiograms (ECG) and central nervous disorders including psychiatric diseases being reported recently. Parisi P et al. presented a family study in which Brugada syndrome, which is characterized by Brugada-ECG and sudden cardiac death, and epilepsy coexist in the same family and this report indicated that familial abnormalities of the central nervous system is reflected on electrocardiograms [6]. In patients with schizophrenia, the Brugada-ECG pattern was seen more frequently [7, 8] and longer QT interval prolongation was observed [9].

However, the percentage of the patients who show Brugada-ECG is not high, and the reported average QT prolongation is not within an abnormal range. Moreover, the associations between these findings and psychiatric clinical factors are not clear. In addition to Brugada-ECG and longer QT interval, early repolarization pattern (ERP) is known as a finding showing abnormal repolarization. ERP and J-point elevation pattern, which are not uncommon and often observed in younger and healthier people [10], are arrestive electrocardiographic patterns associated with sudden cardiac death [11,12]. Recently, it has been reported that there are associations between ERP and attention-deficit hyperactivity disorder [13,14],and the patients with suicidal risks [15]. Moreover, Fitzgerald et.al reported that Patients with serious mental illness, especially schizophrenia/schizoaffective disorder, exhibit a significantly high rate of abnormal late potentials on Signal-averaged electrocardiogram (SAECG) and ERP [16]. However, the prevalence of ERP and the association between ERP and clinical findings are remained unclear.

Moreover, in clinical practice, many patients with schizophrenia are already on medication [7,8], and it is important to consider the effect of these drugs on electrocardiograms. Antipsychotics, mood stabilizers, and antidepressants are known to cause electrocardiogram changes such as QT prolongation and/or Brugada-ECG [17,18]. With regard to ERP, it has been reported that their frequency decreases in multi-antiepileptic combination cases [19] or no significant effect [20] contrary to QT prolongation or Brugada-ECG. To date, the effects of psychiatric drugs on early repolarization pattern have not been reported. The purposes of the present study are follows. First, we retrospectively compared the presence of ERP and J-point elevation patterns between schizophrenic patients and coronary risk factors-matched controls. Coronary risk factor-matched controls were selected from medical checkups. We also used logistic regression analysis in order to investigate the associations between ERP or J-point elevation pattern and confounding factors including psychiatric drug use (caluculated by chlorpromazine equivalent [21] and use of depolarization-blocking drugs [22]). Secondly, we used logistic regression analysis in order to investigate the relationship between clinical findings such as psychiatric family history and electrocardiographic findings among the patients.

## Methods

### Participants

We performed a retrospective age-, sex-, and coronary risk factors-matched case-control study on 85 patients with adult schizophrenia (schizophrenia or schizoaffective disorder) who had been hospitalized in the psychiatric departments of Jikei University from January 2013 to December 2017 to ensure validity of diagnosis, and who underwent ECGs for general screening during follow-up examinations.

Considering that the patients having greater coronary risk factors, controls matching the patients in terms of age, sex, presence of dyslipidemia (DLP), diabetes mellitus (DM), or smoking, were selected randomly from medical checkups of 89 subjects at Jikei University hospital who did not have a diagnosis or a family history of psychiatric disorders within second degree relatives. For diagnoses of schizophrenia and schizoaffective disorder, we followed the criteria of the Diagnostic and Statistical Manual of Mental Disorders, 4th Edition, Text Revision (DSM-IV-TR) (American Psychiatric Association, 2000).

We excluded electrocardiograms in hypothermia or hyperthermia, and patients or controls with structural heart disease, bundle branch block, or WPW syndrome [23]. Additionally, we also excluded ECG contaminated by artifacts or electromyogram noise.

A flow diagram showing the participants and the reasons for their exclusion is presented in Figure 1. This retrospective study was conducted in accordance with the Declaration of Helsinki and approved by the research ethics committee of Jikei University (Approval number: 30-152). *ECG Evaluation*

**Figure 1.**
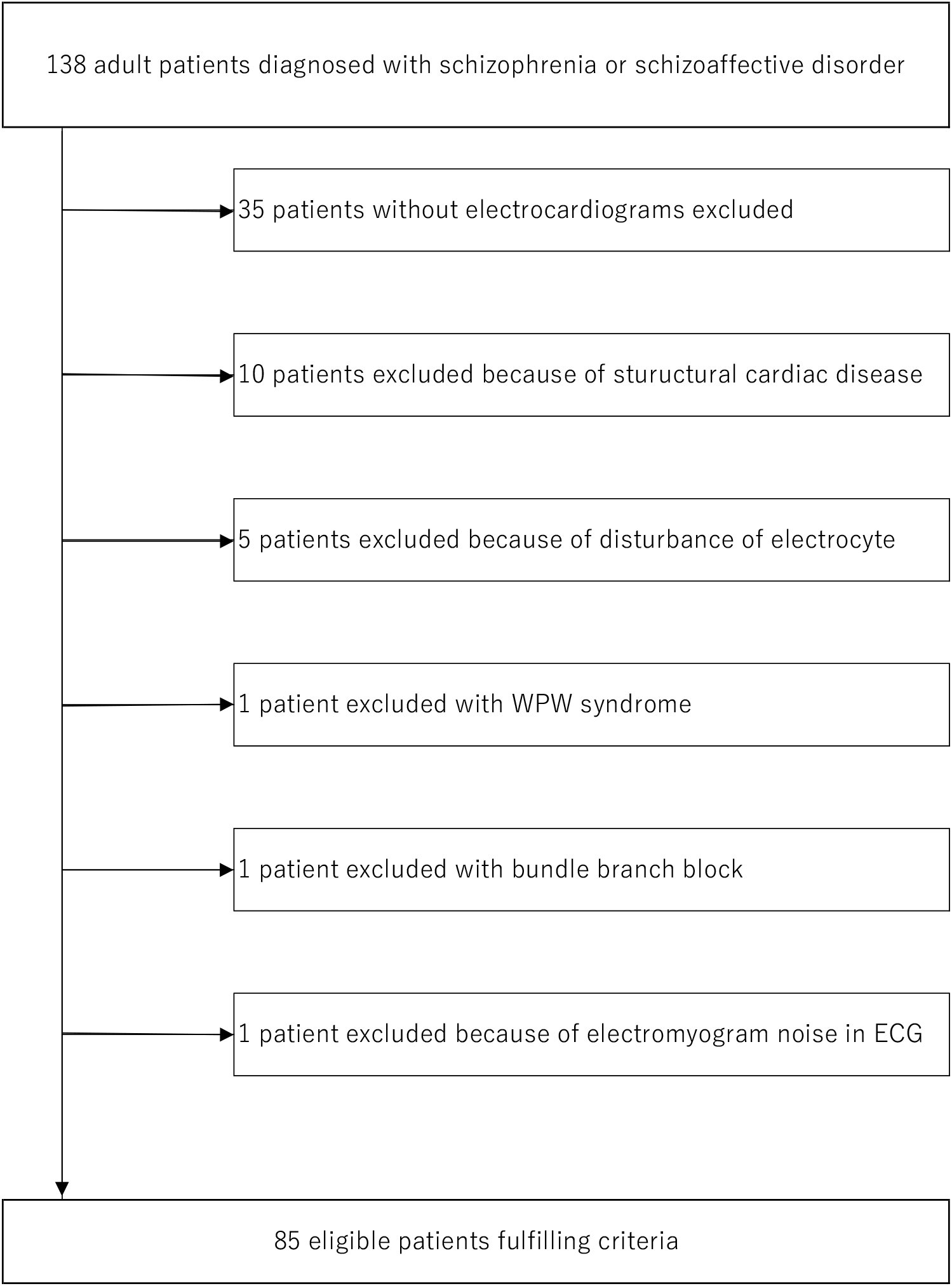
Flow diagram showing study participants.

All ECGs were recorded at a paper speed of 25 mm/sec and amplitude 1 mV/10 mm in the central clinical laboratory or Center for Predicitive Medicine. Two independent reviewers (H.K and K.S) blinded to clinical findings analyzed all the first recorded ECGs without electrolyte disturbance if the results of blood tests were available. If the first ECG was recorded under electrolyte abnormality, the following electrogram was analyzed with normal electrolytes. Disagreements were resolved by consensus. To classify an ECG as ERP or J-point elevation, both reviewers had to be in agreement. We evaluated electrocardiograms in accordance with the method proposed by Peter W. Macfarlane in 2015 [24] as follows. ERP is present if all the following criteria are met. First, there is an end-QRS notch or slur on the downslope of a prominent R-wave. If there is a notch, it should lie entirely above the baseline. The onset of a slur must also be above the baseline. Second, the J-point peak is ≥0.1 mV in two or more contiguous leads of the 12-lead ECG, excluding leads V1 to V3. Third, QRS duration is <120 ms. Both slur and notch should occur in the final 50% of the R-wave downslope to be regarded as ERP in order to exclude fragmented QRS. ST-segment slope should be measured from the termination of the J-point. The ST segment should be regarded as horizontal or downward sloping if the amplitude of the ST-segment 100 ms after termination of the J-point is less than or equal to the amplitude at termination of the J-point. The ST-segment should be regarded as upward sloping if the amplitude of the ST-segment 100 ms after termination of the J-point is greater than the amplitude at termination of the J-point. Additionally, we also evaluated the amplitude of the J-point peak elevation 0.05 mV above the baseline with the criteria met above as J-point elevation pattern. See figure 2 for example of ERP and J-point elevation pattern.

**Figure 2.**
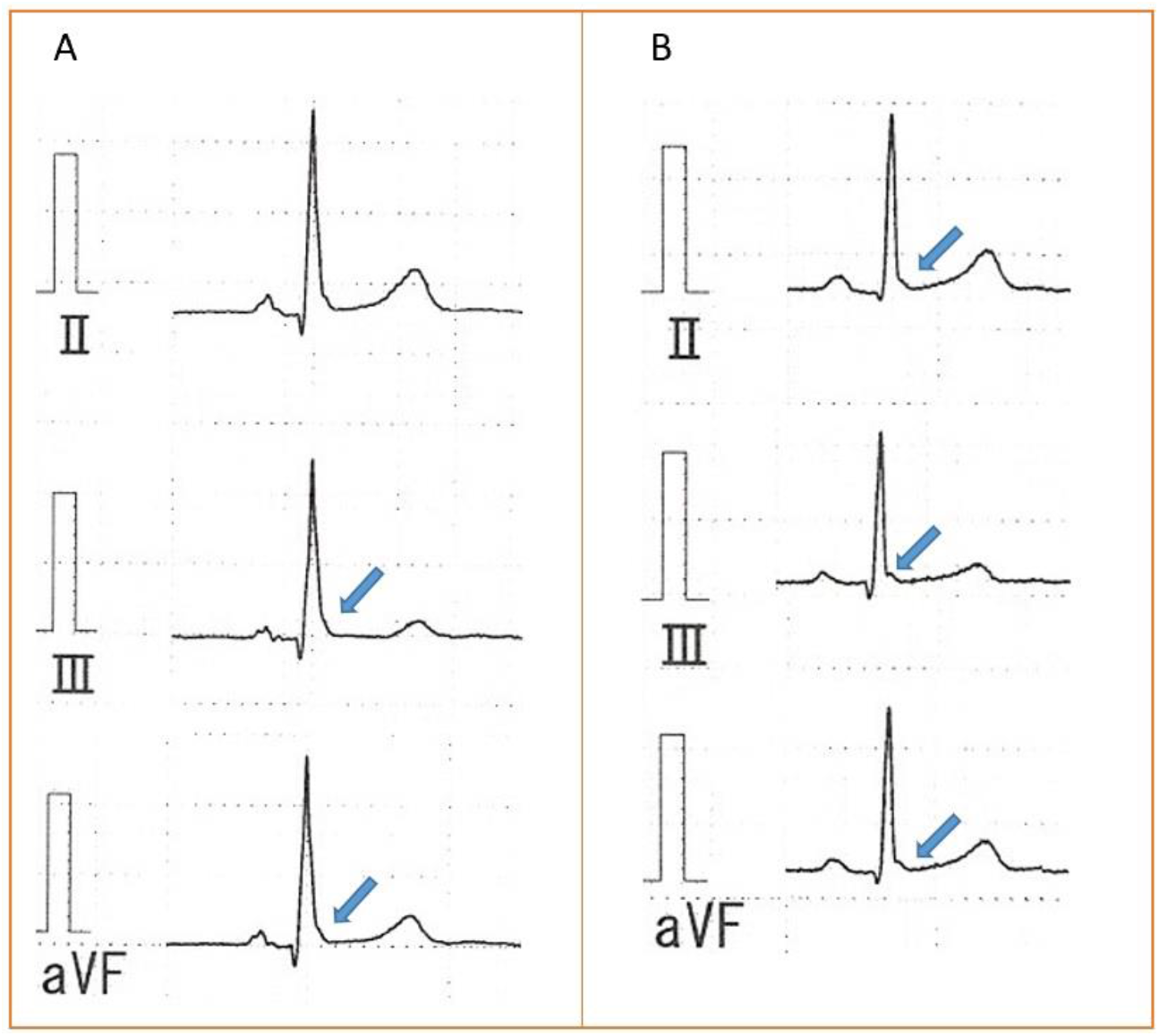
Example of electrocardiograms from schizophrenic patients showing early repolarization pattern in inferior lead (A) and J-point elevation pattern (J-point peak elevation 0.05 mV above the baseline in contiguous leads.) in inferior lead (B).

### Confounders and Clinical Findings

Generally, antipsychotics have been found to influence the human ether-a-go-go related gene (HERG) channel and cause QT prolongation [17]. To account for the effects of antipsychotics medication, an equivalence dose (chlorpromazine equivalent, CPE) was calculated using methods proposed by Inagaki [21]. We also calculated doses equivalent to 100 mg/day of chlorpromazine of newer antipsychotics asenapine equal to olanzapine [25].

Additionally, some psychiatric drugs could affect the J-point and augment Brugada-ECG, which are listed in Brugada Drug’s website [22]. We also evaluated those antipsychotics and mood stabilizers that are listed as depolarization-blocking drugs, which could affect the amplitude of the J point among the patients as previously studied [7,8].

Secondly, we examined the clinical findings of ERP or J-point elevation pattern among the patients. We compared the age, sex and heart rate, and psychiatric family histories of patients within second-degree relatives both with and without ERP or J-point elevation patterns.

### Statistical Analysis

IBM SPSS 25 for Windows (SPSS Japan Inc., Tokyo, Japan) was used for all the statistical analyses. The normality of the continuous variables was tested using the Kolmogorov–Smirnov test. The continuous variables were compared using an unpaired Student’s T-test or the Mann– Whitney U-test when appropriate. Continuous variables with normal distribution were expressed as mean ± SD and continuous variables with non-normal were expressed as the median [25th, 75th percentiles]. The categorical variables were compared using the chi-square analysis or fisher test when appropriate. Logistic regression was performed in order to evaluate the association between schizophrenia and ERP or J-point elevation pattern, and the diagnosis of schizophrenia was entered as the dependent variable, and the presence of ERP or J-point elevation pattern, sex, age heart rate, coronary risk factors, CPE, and use of depolarization-blocking drugs were entered as independent variables among the patients and control subjects. To evaluate the associations between ERP or J-point patterns and clinical findings in the patients, the presence of ERP or J-point elevation patterns were entered as dependent variables, and sex, age, HR, and psychiatric family history were entered into a regression as independent variables. Two-tail P value < 0.05 was statistically significant.

## Results

### Study participants and clinical characteristic

Table 1 presents the clinical and electrocardiographic characteristics of both groups. 59 of the 85 patients were female. 57 of the 89 matched controls were female. Heart rate was significantly faster (P < 0.001) and PR interval was shorter (P<0.001) in the patients’ group. None of the subjects in the control group were under antidepressants, mood stabilizers, or antipsychotics. The presence of ERP or J-point elevation patterns was more prevalent in the schizophrenic group (Both P <0.001). The characteristics of ERP and J-point elevation in each group were compared between the controls and the patients. There was no significant difference between the controls and the patients in ERP amplitude. Both ERP and J-point elevation were more prevalent in the inferior lead in the patients (ERP: P = 0.005, J-point elevation pattern: P = 0.011). Upsloping pattern was more common in patients; however, there was no significant difference in presence of horizontal or downsloping ST segment patterns.

**Table 1.** Comparison of clinical characteristics in schizophrenic patients and controls

|  | Patients (N = 85) | Controls (N = 89) | P value |
| --- | --- | --- | --- |
| Males (%) | 26 ( 31 %) | 32 ( 36% ) | 0.52 |
| Age (years) | 41 [ 33-53 ] | 43 [ 34-54 ] | 0.39 |
| Coronary risk factors |  |  |  |
| Diabetes mellitus (%) | 11 ( 13% ) | 14 ( 16% ) | 0.67 |
| Hypertension (%) | 6 ( 7% ) | 3 ( 3% ) | 0.26 |
| Dyslipidemia (%) | 23 ( 27% ) | 25 ( 28 % ) | 1.00 |
| Smoker (%) | 22 ( 26 %) | 22 ( 25% ) | 0.86 |
| Use of depolarization blocking drugs (%) | 14 ( 16 % ) | 0 ( 0% ) | <0.001 |
| Chlorpromazine equivalent (mg/day) | 575 [ 375-913 ] | 0 [ 0-0 ] | <0.001 |
| Structural heart disease | 0 ( 0% ) | 0 ( 0% ) | 1.00 |
| ECG findings |  |  |  |
| Heart rate (bpm) | 73 [ 66-82 ] | 62 [ 57-70 ] | <0.001 |
| PR intervals (ms) | 147 [ 139-163 ] | 156 [ 147-173 ] | <0.001 |
| QRS duration (ms) | 94 [ 88-99 ] | 95 [ 90-100 ] | 0.277 |
| QTc intervals (ms) | 416 ± 24 | 414 ± 19 | 0.236 |
| ERP (%) | 23 ( 27% ) | 6 ( 7% ) | <0.001 |
| J-point elevation (%) | 34 ( 40% ) | 12 ( 13% ) | <0.001 |
| J point amplitude of ERP (mm) | 1.7 [ 1.3-2.0 ] | 1.6 [ 1.5-1.9 ] | 0.978 |
| ERP locations |  |  |  |
| Inferior leads (%) | 15 (18%) | 4 ( 4% ) | 0.005 |
| Left precordial leads (%) | 4 (5%) | 1 ( 1% ) | 0.20 |
| Lateral leads (%) | 2 (2%) | 0 ( 0% ) | 0.24 |
| Both inferior and lateral leads (%) | 1 (1%) | 1 ( 1% ) | 1.00 |
| ST segment pattern |  |  |  |
| Upsloping pattern (%) | 18 (21%) | 2 ( 2% ) | <0.001 |
| downsloping or horizontal pattern (%) | 4 (5%) | 4 ( 4% ) | 1.00 |
| J-point elevation location |  |  |  |
| Inferior leads (%) | 21 (25%) | 9 ( 10% ) | 0.011 |
| Left precordial leads (%) | 3 (4%) | 1 ( 1% ) | 0.36 |
| Lateral leads (%) | 5 (6%) | 0 ( 0% ) | 0.03 |
| Inferior and lateral leads (%) | 3 (4%) | 2 ( 2% ) | 0.68 |

Data are expressed as numbers, Value as mean ± SD, no.(%) or median [25th, 75th percentiles]. Abbreviations: ERP: early repolarization pattern.

All but three of the patients were under antipsychotic medication. 52 out of 85 patients took second-generation antipsychotic drugs only; five patients took first-generation antipsychotics only. 23 patients took both first-generation antipsychotics and second-generation antipsychotics. 13 patients were prescribed mood stabilizers with depolarization-blocking effects as an adjunctive therapy (Lamotrigine for 7 patients, Carbamazepine for 5 patients, Lithium for 5 patients). No patients were under antiarrhythmic drug use.

### Predictors for ERP or J-point Elevation Patterns among the patients and controls

After multivariable logistic regression between the patients and the sex- and age-matched controls, we found that the diagnosis of schizophrenia is an independent predictor for the presence of ERP and J -point elevation pattern (ERP: P = 0.001, J-point elevation pattern P<0.001). (Table 2). Confounding factors, such as coronary risk factors, resting heart rate, chlorpromazine equivalent and use of depolarization-blocking drugs were not significant predictors.

**Table 2.** Logistic regression among the healthy controls and the patients to predict the presence of early repolarization patterns and J-point elevation patterns

| ERP |  |  |  |  |  |
| --- | --- | --- | --- | --- | --- |
| Variables | B Coefficient | Standard Error | 95%CI of OR |  | P value |
|  |  |  | Lower Bound | Upper Bound |  |
| Schizophrenia (presence) | 2.121 | 0.623 | 2.458 | 28.276 | 0.001 |
| Age (per year) | 0.014 | 0.018 | 0.979 | 1.050 | 0.441 |
| Sex (male) | 0.142 | 0.503 | 0.430 | 3.088 | 0.78 |
| Heart rate (per bpm) | -0.010 | 0.020 | 0.952 | 1.029 | 0.598 |
| DM (presence) | -0.258 | 0.697 | 0.197 | 3.029 | 0.771 |
| HT (presence) | -0.094 | 0.924 | 0.149 | 5.569 | 0.919 |
| DLP (presence) | 0.376 | 0.505 | 0.542 | 3.918 | 0.456 |
| Smoking (presence) | 0.396 | 0.513 | 0.544 | 4.062 | 0.44 |
| Chlorpromazine equivalent ( mg/day ) | -0.001 | 0.001 | 0.998 | 1.000 | 0.262 |
| Depolarization-blocking-drugs (presence) | 0.576 | 0.657 | 0.491 | 6.446 | 0.381 |
| J-point elevation |  |  |  |  |  |
| Variables | B Coefficient | Standard Error | 95%CI of OR |  | P value |
|  |  |  | Lower Bound | Upper Bound |  |
| Schizophrenia (presence) | 1.995 | 0.521 | 2.649 | 20.399 | <0.001 |
| Age (per year) | 0.019 | 0.016 | 0.988 | 1.052 | 0.82 |
| Sex (male) | 0.100 | 0.441 | 0.466 | 2.623 | 0.820 |
| Heart rate (per bpm) | -0.009 | 0.018 | 0.958 | 1.026 | 0.613 |
| DM (presence) | -0.419 | 0.610 | 0.199 | 2.172 | 0.492 |
| HT (presence) | -0.128 | 0.821 | 0.176 | 4.397 | 0.876 |
| DLP (presence) | 0.296 | 0.442 | 0.565 | 3.198 | 0.503 |
| smoking (presence) | 0.260 | 0.455 | 0.532 | 3.162 | 0.568 |
| Chlorpromazine equivalent ( mg/day ) | -0.001 | 0.001 | 0.998 | 1.00 | 0.111 |
| Depolarization-blocking-drugs (presence) | 1.000 | 0.639 | 0.777 | 9.50 | 0.117 |

Abbreviations: CI, confidence interval; DM; diabetes mellitus, DLP, dyslipidemia; HT, hypertension; OR, odd ratio.

### Associations Between Clinical Findings and ERP or J-point Elevation Patterns in Schizophrenic Patients

Familial psychiatric history of schizophrenia is an independent predictor for the presence of ERP, and older age and familial psychiatric history are independent predictors for J-point elevation patterns (Table 3).

**Table 3.** Logistic regression in schizophrenic patients to predict presence or absence of ERP and J-point elevation

| 95%CI of OR |  |  |  |  |  |
| --- | --- | --- | --- | --- | --- |
| Variables | B Coefficient | Standard Error | Lower Bound | Upper Bound | P value |
| Predictor for ERP |  |  |  |  |  |
| Sex (male) | 0.952 | 0.590 | 0.815 | 8.232 | 0.107 |
| Age(year) | 0.032 | 0.02 | 0.994 | 1.073 | 0.098 |
| Heart rate (bpm) | -0.02 | 0.02 | 0.941 | 1.020 | 0.321 |
| Psychiatric family history | 1.519 | 0.551 | 1.552 | 13.45 | 0.006 |
| Predictor for J-point elevation |  |  |  |  |  |
| Sex (male) | 1.059 | 0.545 | 0.990 | 8.394 | 0.052 |
| Age (year) | 0.038 | 0.018 | 1.002 | 1.076 | 0.038 |
| Heart rate (bpm) | -0.028 | 0.019 | 0.936 | 1.01 | 0.143 |
| Psychiatric family history | 1.27 | 0.519 | 1.287 | 9.86 | 0.014 |

Abbreviations: CI, confidence interval; ERP, early repolarization pattern; OR, odd ratio.

## Discussion

In this study, schizophrenia patients in our hospital showed an increased probability of ERP and J-point elevation patterns, and in our schizophrenic patients, familial psychiatric histories were demonstrated to be independent predictors for both patterns.

In previous studies, the prevalence of ERP was characterized by J wave ranges between 1.3 % and 11.2 % in the general Asian population [10, 26]. Considering these results, ERP was more markedly observed in our schizophrenic patients. In our study, however, no patients had Brugada-ECG, which may be due to 70% of the patients being female as well as the small sample size compared with the previous studies [7, 8]. There was no difference in QTc time between the patient group and the control group, which may be due to the higher heart rate in the patient group.

### Possible Mechanism of Early Repolarization Pattern Among Patients With Schizophrenia

In the present study, electrocardiographic findings such as ERP and J-point elevation patterns were significantly more frequent in patients with schizophrenia than among the control group. Several possibilities may explain these results.

Initially, we thought a genetic point of view may explain these results. Mutations in ion channels can be considered gene changes that can cause electrocardiographic changes, and the diseases that are related to these mutations are being studied as cardiac ion channel diseases [27]. The CACNA1C gene is known as a crucial gene in sudden cardiac death associated with ERP [28]. Previous epidemiological surveys have reported that mutations of CACNA1C were associated with psychiatric disorders such as schizophrenia [29]. Considering these previous reports, there might be some genetic overlap between psychiatric disorders and cardiac channelopathies.

As a second hypothesis, it may be considered that electrocardiogram abnormalities were due to chronic inflammation. Chronic inflammation has recently attracted attention as a cause of mental illness. A previous study has shown over-activation of microglia in PET studies in patients with schizophrenia [30]. In another study, examination of peripheral blood also showed elevation of IL-2, IL-6, IL-10, and IFN-γ among patients with schizophrenia [31]. Schizophrenia is a progressive disease and linked to chronic inflammation, which has been mentioned as a possible cause [32]. There are many reports suggesting a relationship between ECG abnormalities and inflammation. It is reported that IL-6 was elevated in subjects with ERP [33]. The frequency of ERP was higher in psoriasis patients compared to controls [34]. These reports suggest that the existence of chronic inflammation is also considered to be a possible factor of early repolarization patterns, as observed in schizophrenic patients.

### Effects of Drugs on ERP

We should also consider the effect of psychiatric drugs on electrocardiograms. Some psychiatric drugs are reported to be associated with drug-induced Brugada-ECG [22]. In addition to these drugs, some psychiatric medications could have depolarization-blocking properties [35, 36]. However, contrary to Brugada-ECG, ERP was reported to be attenuated by depolarization-blocking drugs [37].

Additionally, an antiepileptic drug, which is also used as a mood stabilizer and is partially reported to be a depolarization-blocking drug, did not have a significant effect on ERP [20]. It is even reported that ERP is less likely to be recognized in patients who were under multiple antiepileptic drugs [19].

It is also known that antipsychotics act not only on depolarization but also on repolarization, and the mechanism that mainly affects repolarization is HERG channel-mediated QT prolongation [17]. However, there are no reports on the effects associated with Ito channels [38], which is important in ERP. As far as we know, it has not been reported that the use of antipsychotic drugs has caused manifestations of ERP.

Even drug-induced ERP by sodium channel blockers was thought to be rare [39]. In this study, use of depolarization-blocking drugs or chlorpromazine equivalents were excluded as a significant predictor for ERP or J-point elevation pattern, and these results are compatible with these reports.

Moreover, it has been reported that anti-cholinergic effects attenuated early repolarization pattern [40], and administration of antipsychotics may not contribute to the induction of early repolarization.

In small numbers of patients, ERP was augmented by sodium channel blockers [41, 42]. These patients showed J point elevation in the anterior leads similar to Brugada-ECG. However, in our patients, no patients showed suspected Brugada-ECG patterns.

### Limitations

This study is a retrospective single center study. Therefore, the findings may not be generalizable to the wider population. We did not perform genetic tests associating cardiac ion channels or evaluate for inflammatory markers including CRP. Future studies need to consider these parameters in order to clarify their role in the underlying pathophysiology between schizophrenia and ERP or J-point elevation pattern. The prescribed drugs could contribute to the occurrence of ERP or J-point elevation pattern. Although we adjusted for the effect of the antipsychotic drugs by converting their doses to chlorpromazine equivalents, the effects of drug interaction were not evaluated. The patients or controls without repolarization abnormalities during the daytime may exhibit ERP or J-point elevation pattern at night. Therefore, future studies should consider performing the ECGs both at night and during the day.

## Conclusion

This preliminary case-control study in a predominantly female hospitalized schizophrenic population suggests that there may be associations between ERP or J-point elevation pattern, and this diagnosis and the presence of a family history. Further, more rigorous studies controlling for drug use, hospitalization, and genetic factors should be developed in the future.

## Data Availability

N/A

## Acknowledgments

None.

## Conflict of Interest Statement

None.

## Notes

**Funding:** none

### Competing Interest Statement

The authors have declared no competing interest.

### Author Declarations

This retrospective study was conducted in accordance with the Declaration of Helsinki and approved by the research ethics committee of Jikei University (Approval number: 30-152).

